# Formal educational attainment and HIV treatment adherence in southern and eastern Africa

**DOI:** 10.1101/2025.08.06.25333107

**Authors:** Stephanie Chamberlin

## Abstract

Social policy makers frequently leverage formal schooling as a tool for curbing the HIV epidemic in sub-Saharan Africa. Yet, in the era of ‘Treatment as Prevention’, evidence about the association between formal education and chronic HIV care and treatment in the region remains limited. In this study, I use population-level data from the first round of the Population HIV Impact Assessment to examine the association between years of formal education and HIV treatment adherence (measured via viral load suppression) across seven southern and eastern African countries. Given persistent gender and age disparities in both education and HIV care in the region, I further test for moderation of these associations by gender and age. I find no association between education and viral load suppression in the pooled regional sample (N=12,198), in country-specific analyses, and no modification of these findings by gender or age. Further, results were robust in sensitivity analyses using different measures of educational attainment. These somewhat surprising findings challenge our common understanding about education as a catalyst for improved health, and provide theoretical insights into what may drive the relationship (or lack thereof) between education and chronic health in different contexts. More research is needed into the contextual factors and countervailing mechanisms that may explain such results.

## Introduction

Education—specifically formal schooling— has long been imagined as a powerful tool to address the HIV epidemic in sub-Saharan Africa. Existing evidence is almost entirely focused on the role of schooling in reducing sexual risk behaviors, particularly among those who are uninfected (1, 2). Yet, in the current era of ‘treatment as prevention’, we know remarkably little about how formal education can support HIV treatment adherence among people living with HIV (PLWH) (3, 4). Those PLWH who take HIV treatment regularly, leading to viral suppression, have nearly zero risk of transmitting the virus sexually and are more likely to lead long and healthy lives (5). Thus, increasing viral load suppression (VLS)—a common biomarker for HIV treatment adherence—among PLWH is a key public health objective for addressing the dual nature of HIV as both a communicable and chronic condition. Despite dramatic improvements in HIV treatment access under Universal Treatment policies in southern and eastern Africa since 2010, improvements in HIV outcomes have been unevenly distributed (4) (6, 7). By examining educational disparities in viral load suppression, this study is responsive to calls to better understand and respond to factors that contribute to persistent disparities in HIV care and treatment in southern and eastern Africa(6, 8), the region of the world with the highest HIV prevalence.

As one of the most widely diagnosed and treated chronic conditions in sub-Saharan Africa, the case of HIV also provides a unique opportunity to study educational associations with chronic care more broadly at the population level. In a context where the prevalence of chronic health conditions is rapidly increasing, many in the global health community are now looking to the successful expansion of HIV treatment in the sub-Saharan African region as an example for expanding chronic care for other health issues (9). Indeed, chronic HIV treatment mirrors numerous other conditions that require daily or routine medication and routine medical visits over the course of one’s life to avoid life altering consequences. For most chronic conditions, care and treatment is only recently and slowly expanding in sub-Saharan Africa (10), which has historically hampered our ability to study regional associations between education and chronic care management (11). By leveraging insights from the case of HIV, this study may inform more equitable chronic care interventions in the region.

Moreover, this research expands our theoretical and empirical understanding of education-health associations across different conditions and contexts. Within sub-Saharan Africa, inconsistent findings from prior research introduce important questions about potential differences in the association between education and HIV treatment adherence across countries (3, 12). It remains unclear the extent to which inconsistencies in education-HIV treatment associations reflect measurement differences versus true contextual variation. I address this by using consistent measures available from the Population HIV Impact Assessment (PHIA) to examine the relationship between formal education and viral load suppression (a key indicator of treatment adherence) within and across seven southern and eastern African countries. Further, given well-documented age and gender disparities in HIV outcomes and schooling in the region (13–15), I study the potential for differences in these associations by gender and age or cohort.

This work provides a novel perspective about the applicability of previous theoretical frameworks for different types of health issues and across geographic settings. Within sub-Saharan Africa, our current knowledge about positive education-health associations is largely limited to maternal and child health outcomes among young women of reproductive age (16–19). In contrast, I focus on a chronic health outcome among adult men and women of all ages, in diverse contexts across southern and eastern Africa. Many factors that may have explained educational associations with maternal and child health (e.g., access to one-off clinic visits or nutrition) are likely different for the on-going management of a chronic condition (12). Much theoretical work about the associations between education and health has been developed using insights from the global north. Those socio-economic factors (e.g., employment opportunities) that may explain educational associations with chronic conditions in higher-income countries, like the U.S. and Western Europe, may not hold in the southern and eastern African context (20). Through the research presented here, I provide a foundation for theoretical debate and refinement that incorporates formal schooling and chronic health in the global south.

## Background

### Education and HIV Treatment Adherence

There are numerous reasons to believe that education could have a positive influence on chronic HIV treatment in sub-Saharan Africa. First, education may increase an adult’s employment and wealth, in turn providing individuals with material resources that can be used to overcome logistical barriers to chronic care services (21). For example, for people to effectively adhere to their HIV treatment (aka, antiretroviral therapy (ART or ARVs)) they must return to the health clinic on a monthly or quarterly basis to obtain their medication and be monitored for medication effectiveness. These ongoing clinic visits necessitate money for transportation and the ability to negotiate and compensate for time off of work (22, 23). Expanded access to HIV treatment in the region means that these routine visits can now be carried out at local clinics. However, even local clinic visits can involve travel of an hour or more and long wait times (24). To the extent that more education provides greater access to financial resources, transportation, and/or more flexible work arrangements, more educated PLWH may find these financial and time costs easier to manage.

Second, education can facilitate the development of key skills (e.g., numeracy and literacy), motivation, and self-efficacy that can positively influence an individual’s treatment adherence (25–27). First, those who can read and write may be more effective in maintaining their HIV care and treatment. Appointment dates are often written down in a client’s personal health record (i.e., health passport) that necessitates literacy skills to remember appointments (28, 29). Other tools for remembering appointments, like calendars and cell phones, additionally require basic literacy skills. Second, education may be protective, as more educated clients may be more inclined to advocate for themselves due to greater self-efficacy and feel more empowered to return to care rapidly after a missed appointment (30). Further, those with more education may more accurately assess the risk of treatment non-adherence and have greater motivation to maintain HIV care and treatment, even in the face of other obstacles (31).

Although conventional wisdom and prior literature generally suggest a positive relationship between education and treatment adherence, there are also several reasons to expect the relationship in the sub-Saharan African context may be attenuated. First, limited employment opportunities in many areas result in short supply of financial resources for individuals across the education spectrum (32). Second, despite remarkable gains in accessing schooling, the quality of education has not seen commensurate gains, resulting in limited literacy and numeracy skills gained from attending school (33). Thus, even with more education, individuals may not possess the material or cognitive resources needed to support their HIV treatment adherence. Third, because HIV treatment has become increasingly simple and widely accessible—it is now free and requires just one pill a day—it is plausible that education could play a less important role in treatment adherence than would be expected for more expensive or complex chronic care regimens, or where treatment access is more limited (34).

In some cases education may be *negatively* associated with HIV treatment adherence. Examples from other areas of the world, such as vaccine uptake in the US, show that the more educated may feel more empowered to disregard or tailor medical advice to suit their beliefs or lifestyle (35, 36). Further, prior research in the U.S. and Malawi demonstrates that increased employment demands may actually create a barrier to treatment adherence (22, 37). To the extent that education is associated with greater and more demanding types of employment among the more educated, this may result in greater barriers to routine treatment adherence. Even when education does confer some positive benefits for viral load suppression, the presence of such negative effects—even if not universal—may attenuate positive population-level associations.

### Gender differences

Gender differences in education, health care access, and power dynamics suggest that the association between education and viral load suppression may be distinct for men and women. In this study, I move beyond well-known gender disparities in both education and HIV care (13, 38), to examine how educational attainment may benefit men’s and women’s chronic HIV treatment in different ways.

Men’s greater access to employment and income suggests that the benefits of education for HIV treatment adherence may be more pronounced for men. Even when women obtain higher levels of education, they are less likely to work outside of the home and earn less money when compared to similarly educated men (39). Thus, it is not clear that having more education confers a substantially greater material advantage to women—for example, for managing frequent trips to the HIV clinic. In contrast, more educated men may have comparatively greater material resources to facilitate their routine HIV care than less educated men (32).

Men and women may exhibit different associations between education and HIV treatment adherence as a consequence of their different levels of comfort with using health services in general and historically different experiences with accessing HIV care. Health care systems in sub-Saharan Africa have largely centered on women of reproductive age, with a focus on antenatal care services and under-five vaccinations and check-ups (40). Women were the first recipients of universal HIV treatment for life, at least five years before universal treatment was expanded for men (40). As such, women across the education spectrum are likely to have more experience and comfort with managing their HIV treatment. Whereas men with more education may have had greater opportunities to seek HIV treatment in private clinics prior to the expansion of universal HIV treatment, and may generally have greater self-efficacy and experience seeking health services than their less educated counterparts. These gendered dynamics pertaining to financial resources and comfort attending health clinics suggest that positive education-viral load suppression associations will be stronger for men and weaker for women.

On the other hand, HIV treatment adherence is not only a matter of clinic access—it must also be managed inside the home, where gender dynamics are particularly entrenched. More educated women may be better equipped to negotiate gendered power dynamics and stigma in the home, which are frequent barriers to women’s treatment adherence and HIV clinic attendance For example, women in the region report hiding their ART from other household members More educated women likely face less precarity due to HIV stigma for a variety of reasons, including greater financial resources outside of marriage, wider social networks, and their individual skill sets (43). This suggests the alternative possibility that there will be a strong positive association between education and viral load suppression for both men and women.

### Age differences

Across the life spectrum, it is plausible that education could be a resource to help individuals navigate a variety of life obstacles that impede HIV care and treatment. There are well documented differences in HIV outcomes by age in the region (14), with worse outcomes among younger adults, suggesting that older and younger individuals may experience different barriers to HIV care. Life demands such as work and child rearing are common in younger adult populations, which likely create competing priorities for the demands of on-going HIV care and treatment. Older individuals may have other complications such as greater mobility issues and more chronic health concerns that complicate their HIV treatment adherence. While each set of challenges are unique, it is likely that education could be an important resource for overcoming the HIV treatment barriers faced by both older and younger populations—suggesting positive education-viral load suppression associations for younger and older adults alike.

On the other hand, a cohort perspective suggests that people of different ages likely obtained different sets of resources from their schooling, influencing the extent to which their education supports HIV treatment adherence. Given extensive educational reforms in the region over the past 40 years, those in older age groups enjoyed less access to education, but those who did access education may have had better quality schooling (44). As a consequence, older adults with more education are likely to have gained more income-earning opportunities from their schooling credentials and more skills for managing their HIV treatment adherence. Thus, being more educated in an older age group is potentially more differentiating than it might be within younger age groups, for whom attending school was more normative but also lower quality (45). This suggests that in older age groups, those with more education will be better able to adhere to their HIV treatment, but that education will be less differentiating for younger age groups.

The aim of this study is to examine whether education is associated with viral load suppression (as an indicator for HIV treatment adherence) in southern and eastern Africa and whether this association varies by gender and age. Because the theoretical and empirical literature suggests competing hypotheses for the existence or direction of these associations, I do not offer specific hypotheses. I will examine and discuss how my results contrast or support prior theoretical suppositions and empirical information.

## Data and Methods

### Ethics Statement

To conduct this study I used secondary, publicly available Population HIV Impact Assessment (PHIA) data, and I carefully reviewed and followed all documentation and guidelines for appropriately managing and analyzing PHIA data. All PHIA survey protocols, consent forms, screening forms, refusal forms, referral forms, recruitment materials and questionnaires were reviewed and approved by in-country ethics and regulatory bodies and the institutional review boards of Columbia University Medical Center, Westat, and the U.S. Centers for Disease Control and Prevention. All adults provided written or verbal informed consent that was recorded on electronic tablets before beginning the survey. All respondents were informed of the voluntary and confidential nature of the survey and separate consent was obtained for laboratory samples. All publicly available data is free of identifiers and numerous measures are in place to protect respondent confidentiality. PHIA ethical protocols, including procedures to ensure privacy and confidentiality, are outlined in detail in country Final Reports that can be found here: https://phia.icap.columbia.edu/resources/. The data for all seven countries included in this study were accessed by the author upon approval from the PHIA study team, and were downloaded in August of 2022. All data is publicly available upon request from the PHIA study at https://phia-data.icap.columbia.edu/. All statistical code is available upon request from the author.

### Data

To answer the question of whether education is associated with viral load suppression, I draw on the first round of data collection (cross-sectional) from the Population HIV Impact Assessment (PHIA) study (46). I focus on seven countries in southern and eastern Africa for which data collection began between 2015 and 2017, was completed by July 2018, and for which the data was made publicly available by June 2021: Lesotho, Malawi, Namibia, Tanzania, Uganda, Zambia, and Zimbabwe.

### Study Sample

My final analytic sample included 12,198 PLWH with complete data who had initiated HIV treatment by the time of the survey, as documented either via self-report or by evidence of ART in survey blood tests.

The PHIA samples are designed to be population-representative for men and women, ages 15-64, following a two-step process (46). First, enumeration areas were selected with probability proportional to size. Second, households within selected enumeration areas were selected at random. All eligible adults who lived in or slept in each selected house the prior night were eligible to participate. *Household* data include information on household resources/wealth and number of household members; *individual* data include education, age, and information to construct dependent variables, including self-reported engagement with HIV services. B*io-marker* data include laboratory test results for HIV diagnosis, viral load, immune system health (e.g., CD4 count), the presence of ART in the body, and ART resistance. Additional information on the PHIA study design can be found here: https://phia-data.icap.columbia.edu/.

Among the seven countries included in my analysis, there were 17,187 people living with HIV(PLWH) before limiting the analysis to those PLWH who had initiated ART. Among the full sample of PLWH, I excluded 778 respondents from my analyses who had incomplete data on self-reported variables (apart from education) and bio-marker values that were included in my multivariable tests (<1% of my N) or outliers who reported >26 years of education. For those 5% of cases who were only missing data on the key independent variable of education, I imputed the years of education using a single imputation nearest neighbor approach (47).

### Measures

I obtained information on socio-demographic information, including years of education, from linked household and individual datasets. I then linked this information with bio-marker datasets, which contained information on viral load and the presence of ART in the body.

I used the internationally agreed upon limit of a viral load of < 1000 copies/mL (48) to create a dichotomous measure of viral suppression, conditional on having started ART. Viral loads were obtained via blood tests collected as a routine component of the PHIA study among willing participants. 1000 copies/mL is the minimum consistently detectable viral load count available via most viral load lab tests in sub-Saharan Africa, and there is very low risk of disease progression or onward transmission below this level (49). Thus, this is the most clinically relevant cut-off used in health care practice and research throughout the region.

I combined two measures of self-reported education—‘highest level of school you attended’ (e.g., primary, secondary) and ‘highest grade you completed at that level’ (i.e., years of schooling completed at that level) to create one continuous measure for years of education completed—a common practice for population-level surveys from the region (50). For example, someone who reported completing four grades at the secondary level would have a total of 12 years of education assuming primary schooling ends with the completion of grade 8. Because the number of grades in each level and the number of levels of education varied between countries, I used country-specific calculations.

My use of a continuous measure of education contrasts with other commonly used ordinal or categorical measures of primary, secondary, and tertiary level attendance and completion. A continuous measure provides an opportunity to assess the value of education independent of the credentialing offered by completing a given level of education (51), while preserving the variation within a given level of schooling. Further, using a continuous measure of education allowed me to explore any potential curvilinearity in observed relationships. For consistency in comparing my findings with other studies, I provide sensitivity analyses using the different categorical measures of education levels (i.e., none, some primary, completed primary, some secondary, completed secondary, and tertiary) (see S1 Table).

I included individual-level covariates in my multivariable regression models to adjust for key socio-demographic characteristics. I measured age in years using a continuous measure centered at the minimum age for 15, which facilitates the interpretation of the coefficients. I use a self-reported variable for gender as binary measure (men vs. women). Given the high rate of prematurely leaving school and repeating grades in sub-Saharan Africa, I controlled for attendance in school among the adult sample, using a binary measure of *in vs. out of school*. I used an integer measure to control the number of children living in the home, which may suggest differential care-taking and resource burdens. To account for intra-household social support and gender dynamics, I adjusted for a mutually exclusive, categorical measure of marriage: never married, currently in a relationship (married/partnered/co-habitating), and no longer in a relationship (widowed, divorced, separated). To account for other measures of socio-economic status, I used a binary measure of *employed vs. not employed* in the past year, and a five-level index measure of household wealth—from poorest to richest—created by the PHIA study using a principle components analysis and relative quintile breakdown (46). I accounted for geographic differences by controlling for a binary measure of *urban vs. rural*. Finally, I adjusted for key socio-political and -economic differences between countries (e.g., GDP, literacy level, economic development, etc.) —which can influence education and health care access and outcomes—by including country-level fixed effects as dummy variables for each of the seven countries in my models with all countries pooled together.

### Analysis

I conducted all analyses using Stata 18 (52). Analyses include descriptive statistics along with bivariate and multivariable logistic regression models. I applied the individual-level base weights for blood test participation (the level of measurement with the lowest level of participation). All weights were calculated by PHIA to account for individual selection in the PHIA sampling process, which includes adjustments for non-response, the multi-stage stratification process, and under-coverage by age and gender (46). Following guidance from PHIA, I applied weights using jack-knife replication, including appropriately weighting data when pooling across multiple countries (46).

I developed two multivariable logistic models. Model 1 included all covariates listed above, with the exclusion of the potentially mediating socio-economic variables of employment and wealth. Subsequently, in Model 2, I tested the unique influence of education on viral load suppression, net of the of wealth or employment, including all other covariates from Model 1 (Model 2). This allowed me to assess the association between education and viral load suppression, with (Model 2) and without (Model 1) controlling for the influence of education and wealth—providing insight about the value of education beyond material resources— while still controlling for other covariates.

I ran the final Models 1 and 2 individually for each country and for the pooled sample with all countries combined (with country-level fixed effects). Country-specific findings for Model 2 are provided in S2 Table. For Model 3, I tested whether the relationship between education and viral load suppression is moderated by gender by including an interaction term between the binary measure of gender and education. Country-specific findings for Model 3 are provided in S3 Table. For Model 4, I tested whether age moderated the relationship between education and viral load suppression by including interactions between the continuous measure of age and years of education. Country-specific findings are provided in S4 Table. Finally, I adapted Model 2 using a different categorical education measure—entry into or completion of schooling levels (i.e., primary, secondary, tertiary)— that align with some prior research in sub-Saharan Africa (see S1 Table).

## Results

Table 1 provides a breakdown of the sample by country and the weight-adjusted sample for each country within the pooled analysis. As shown in Table 2, of the 12,198 people who started ART, 86.3% were virally suppressed in the pooled data—substantially lower than the internationally recognized goal of 95% viral load suppression. There is no association between education and viral load suppression (OR=0.99, CI (0.97, 1.01)) (Table 2) in unadjusted analyses. There are clear differences in viral load suppression across other socio-demographic characteristics—those who were older, women, and previously or currently in a relationship were more likely to be virally suppressed.

**Table 1.** Analytic sample of PLWH who had initiated antiretroviral therapy, by country.

| Country | Total N (%),<br>unweighted | Weighted % of<br>pooled sample<br>in each country |
| --- | --- | --- |
| Lesotho | 2454 (20%) | 6% |
| Malawi | 1584 (13%) | 16% |
| Namibia | 2070 (17%) | 4% |
| Tanzania | 1007 (8%) | 21% |
| Uganda | 1195(10%) | 20% |
| Zambia | 1433 (12%) | 14% |
| Zimbabwe | 2455 (20%) | 20% |
| **Total (N)** | **12, 198** | **100%** |

**Table 2.** Descriptive characteristics and unadjusted associations with viral load suppression (viral load suppression) among people living with HIV who initiated antiretroviral therapy, odds ratios (OR) with 95% confidence intervals (CI)

|  | No viral<br>load<br>suppression | viral load<br>suppression | OR (CI) |
| --- | --- | --- | --- |
|  | *Mean (SD)* |  |  |
| **Age (years, centered at 15)** | 35.5 (8.2) | 39.4 (10.8) | 1.03 (1.02, 1.03) |
| **Education (years)** | 7.1 (3.4) | 6.8 (3.9) | 0.99 (0.97, 1.01) |
| **Children in home (#)** | 2.2 (2.0) | 2.2 (1.8) | 1.01 (0.97, 1.05) |
|  | *%* |  |  |
| **Gender (%)** |  |  |  |
| Male | 39.0 | 32.9 | (ref) |
| Female | 61.0 | 67.1 | 1.31 (1.12, 1.54) |
| **Relationship status (%)** |  |  |  |
| Never married/partnered | 13.0 | 9.7 | (ref) |
| Previously married/partnered | 30.6 | 31.9 | 1.39 (1.10, 1.76) |
| Currently married/partnered | 69.4 | 68.1 | 1.40 (1.08, 1.80) |
| **Rurality (%)** |  |  |  |
| Rural home | 58.6 | 58.8 | (ref) |
| Urban home | 41.4 | 41.2 | 1.01 (0.85, 1.19) |
| **Current school status** |  |  |  |
| Not in school | 94.5 | 95.8 | (ref) |
| In school | 5.5 | 4.2 | 0.75 (0.56, 1.01) |
| **Employment in past year (%)** |  |  |  |
| Not employed | 42.3 | 42.7 | (ref) |
| Employed | 57.7 | 57.3 | 1.02 (0.87, 1.19) |
| **Wealth quintile (%)** |  |  |  |
| Lowest wealth | 17.5 | 14.9 | (ref) |
| Low wealth | 16.3 | 17.4 | 1.26 (0.96, 1.65) |
| Medium wealth | 20.8 | 21.5 | 1.22 (0.95, 1.57) |
| High wealth | 24.8 | 24.0 | 1.14 (0.89, 1.46) |
| Highest wealth | 20.6 | 22.2 | 1.27 (0.97, 1.66) |
| **Total** | **13.8%** | **86.3%** | **12,198** |

### Education and viral load suppression associations

In Table 3, I provide the estimates from multivariable models testing the associations between education and viral load suppression in the pooled sample. Model 1 does not control for employment or wealth, whereas Model 2 does adjust for these factors. In both models, for each additional year of education there was no difference in the likelihood of achieving viral load suppression—with statistically and practically small effect sizes and narrow 95% confidence intervals (CI) that crossed 1.00 (Model 1: OR=1.01, 95% CI=0.99,1.03; Model 2: OR=1.00, 95% CI=0.98, 1.03).

**Table 3.**
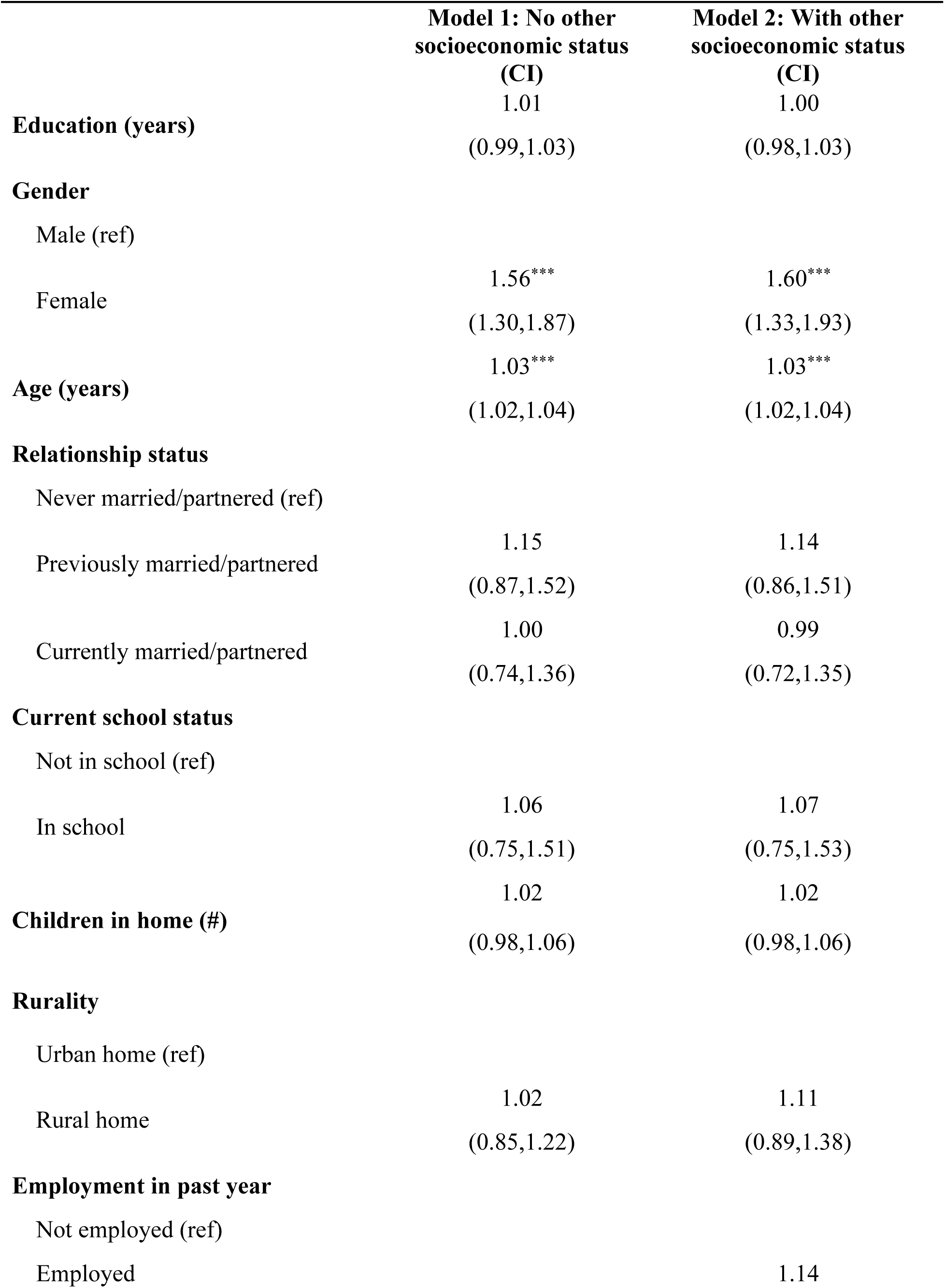
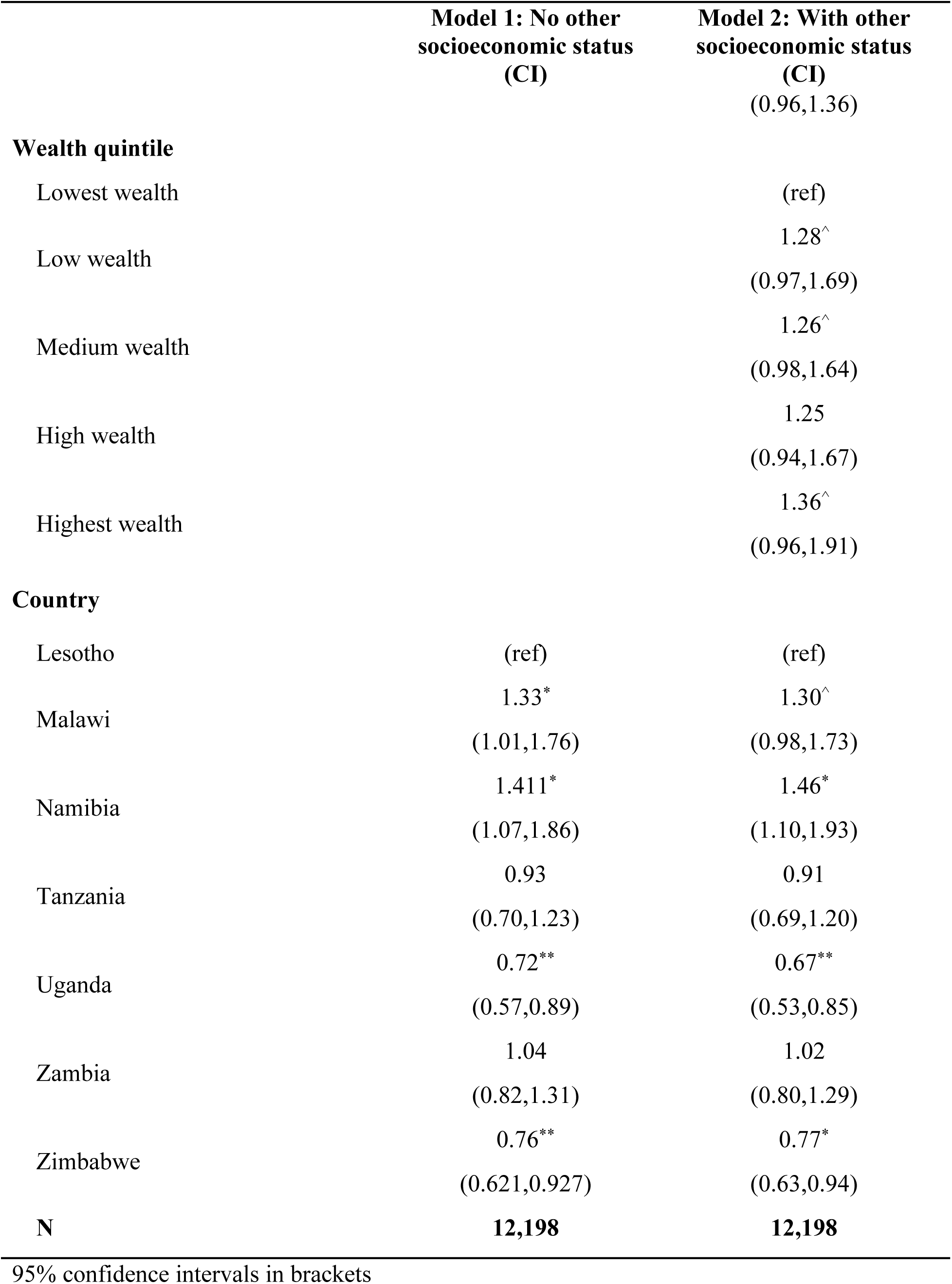
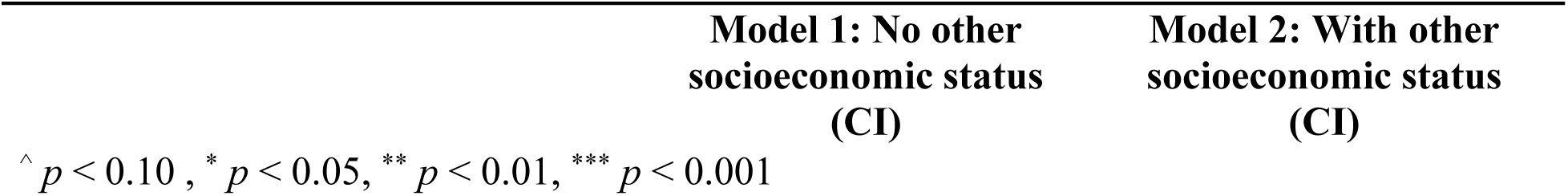
Associations between years of education and viral load suppression (viral load suppression) among people living with HIV who initiated antiretroviral therapy, adjusted odds ratios (AOR) with 95% confidence intervals (CI)

Women were approximately 60% more likely to be virally suppressed comparted men, and for each additional year of age, there was a 3% greater odds of being virally suppressed in both models, controlling for other factors. In Model 2, neither employment nor wealth were associated with viral load suppression when adjusting for other variables.

As shown in Figure 1, the lack of association between education and viral load suppression persisted across all countries. The lack of statistical association between education and viral load suppression was robust across sensitivity analyses—I observed similar findings when I used an ordinal measure of education level (e.g. primary, secondary, tertiary), a binary measure of any vs. no education instead of years of education when included all PLWH regardless of their ART initiation.

**Figure 1.**
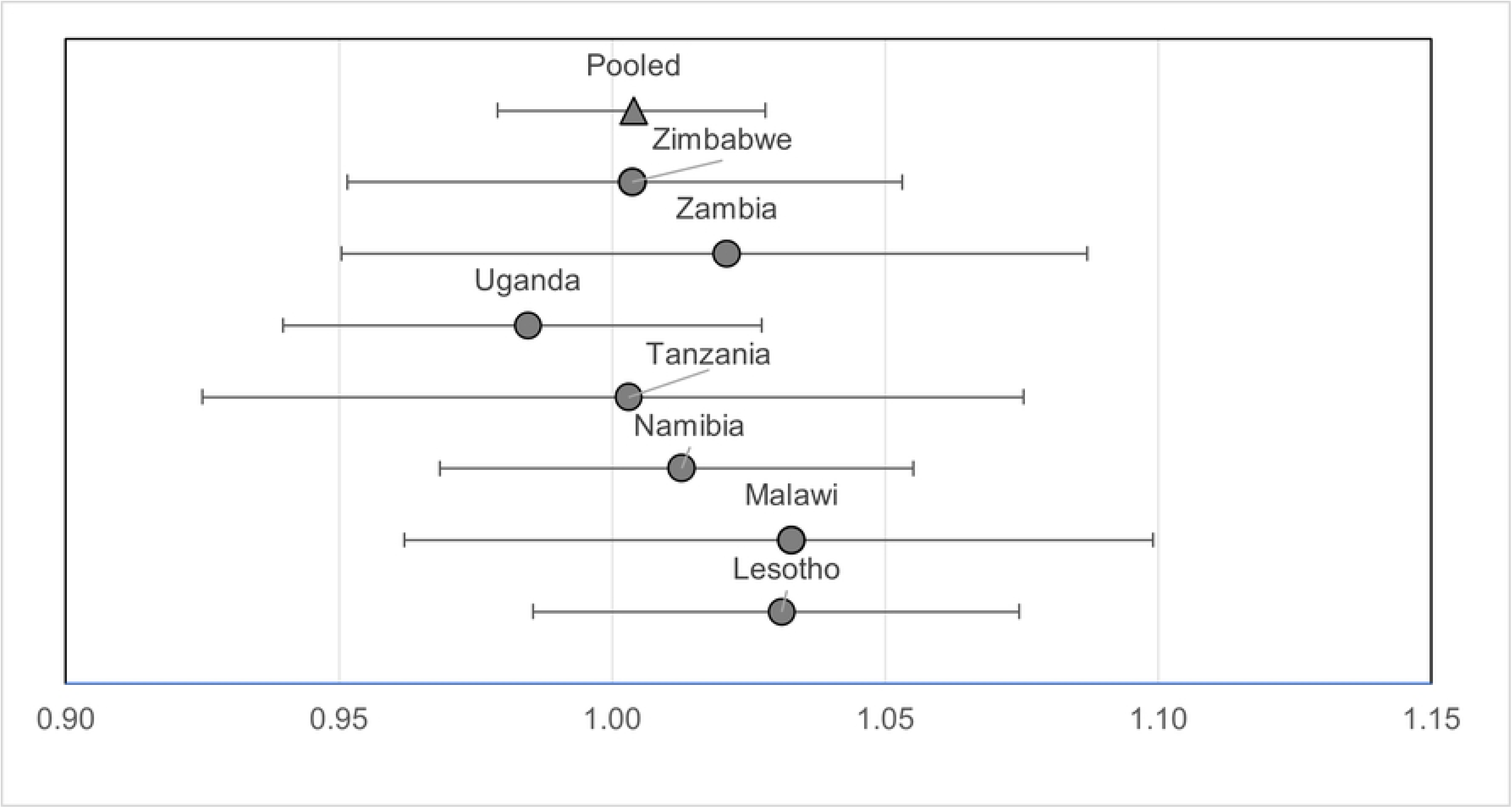
Adjusted odds ratios of viral load suppression for each additional year of individual education among people living with HIV who initiated antiretroviral therapy, with 95% confidence intervals (from Table 3, Model 2).

### Gender moderation results

In Table 4, Model 3 demonstrates that the interaction between gender and education was statistically insignificant at the 95% confidence level, suggesting that there was no moderation in education-viral load suppression associations by gender. Figure 2 more clearly demonstrates the lack of any education-viral load suppression association for both men and women, showing the probability of viral load suppression across all years of education in the pooled model, with two separate lines for men and women. Across the education spectrum, the probability of viral load suppression for men was between 80% and 85%, whereas for women it was between 85% and 90%. In my country-specific sensitivity analyses, gender did not modify educational associations with viral load suppression in any country.

**Table 4.**
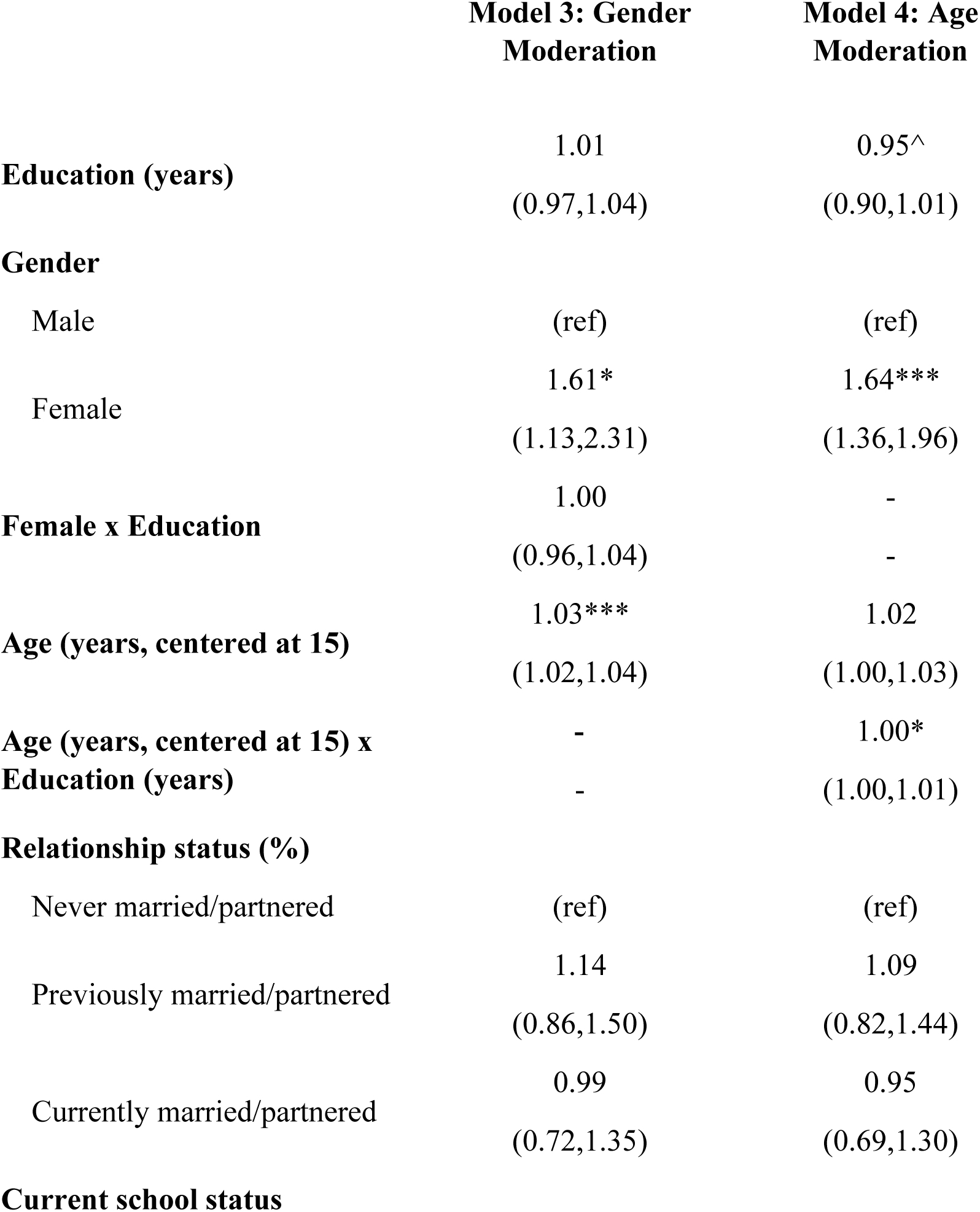
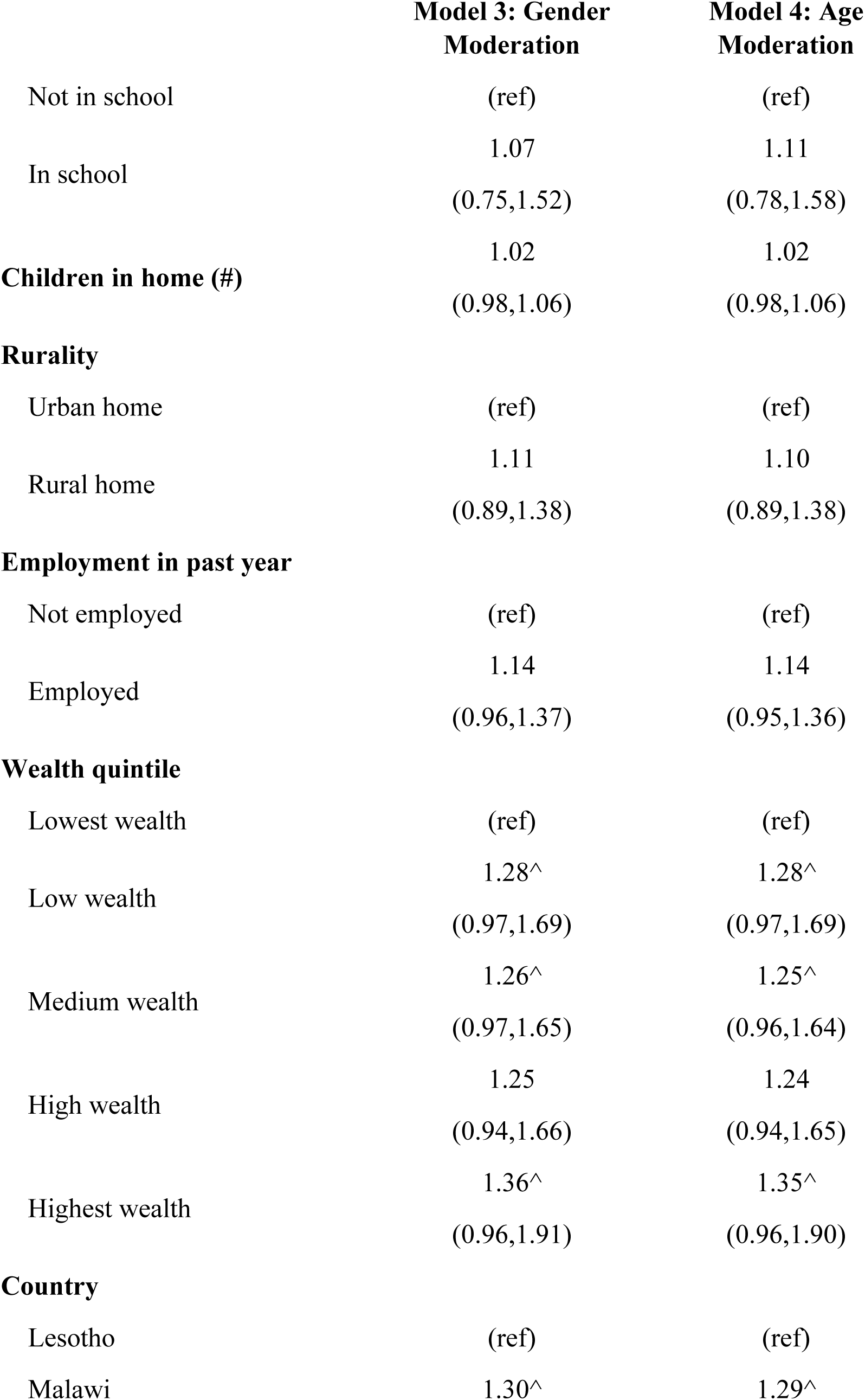
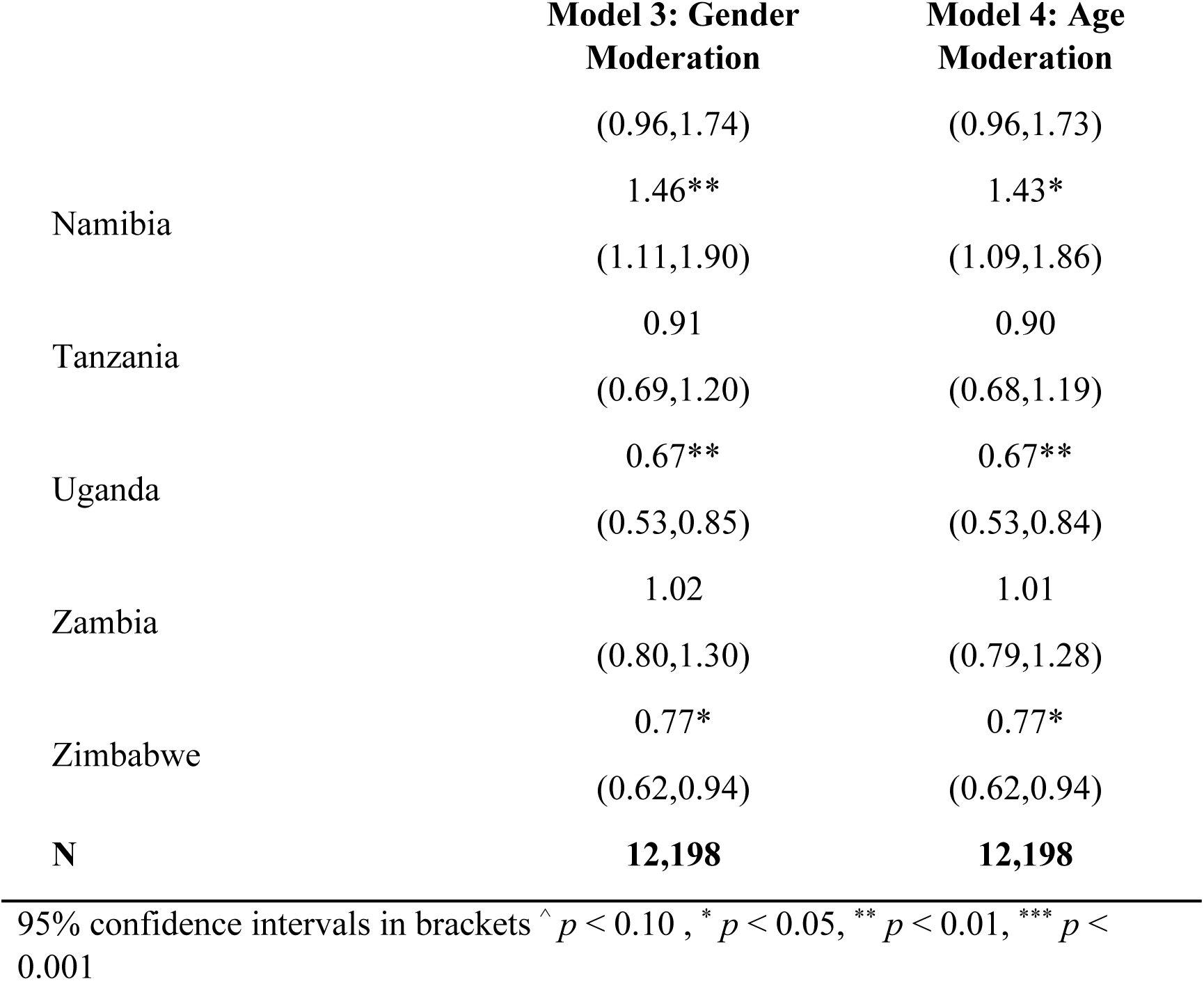
Gender and age moderation of associations between years of education and viral load suppression (viral load suppression) among people living with HIV who initiated antiretroviral therapy, adjusted odds ratios (AOR) with 95% confidence intervals (CI)

**Figure 2.**
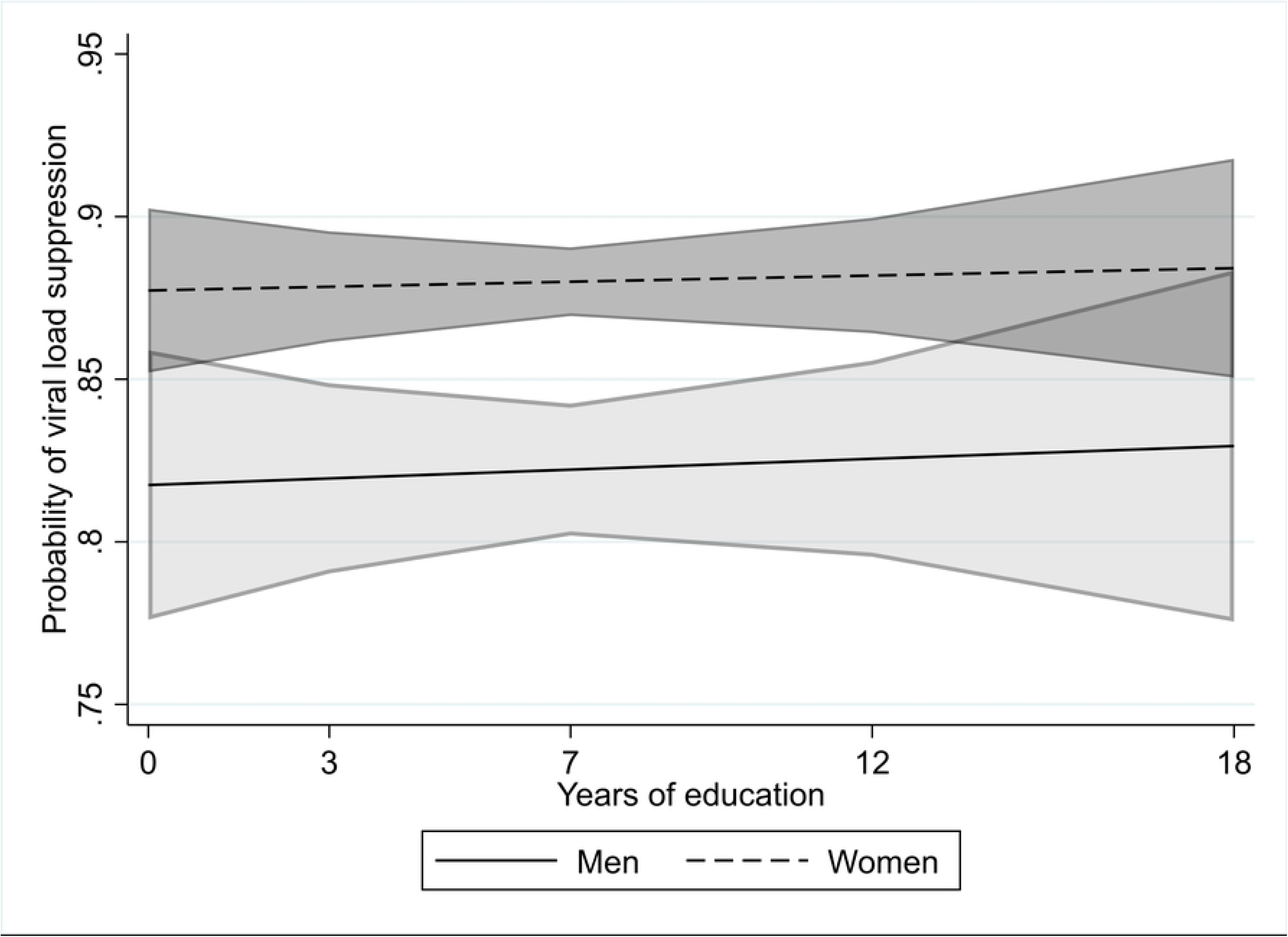
Probability of viral load suppression for each year of individual education with 95% confidence intervals, for men and women living with HIV who initiated antiretroviral therapy (from Table 4, Model 3).

### Age moderation results

In Model 4, the interaction term between education and age was statistically significant at the 95% confidence level, suggesting that age moderates the relationship between education and viral load suppression (see Table 4). For each additional year of education there was a 5% decrease in the odds of viral load suppression when controlling for other factors, but the odds of viral load suppression in relation to education increase by only .2% for each 1-year increase in age. These relationships are more clearly visualized in Figure 3, which displays the probability of viral load suppression across all years of education, with each line representing a different age from 25-60 in the pooled model. Among the 60-year-old age group, there was a positive association; among middle aged groups, there was a statistically significant (at the 95% confidence level) positive association at age 45—reflecting a difference between those with no education vs. those who had 12 or more years of schooling--and there was no association at age 35; and among young adults (age 25) there was a statistically significant negative association (at the 95% confidence level), which reflects a lower probability of diagnosis between for those with no schooling vs. those with 12 or more years of schooling. In sensitivity analyses, these patterns were consistent for both men and women in the pooled sample. However, the education-age interaction term was statistically not significant at the 95% confidence level in all country-specific models, suggesting that age moderation is only detectable in the larger, pooled sample size.

**Figure 3.**
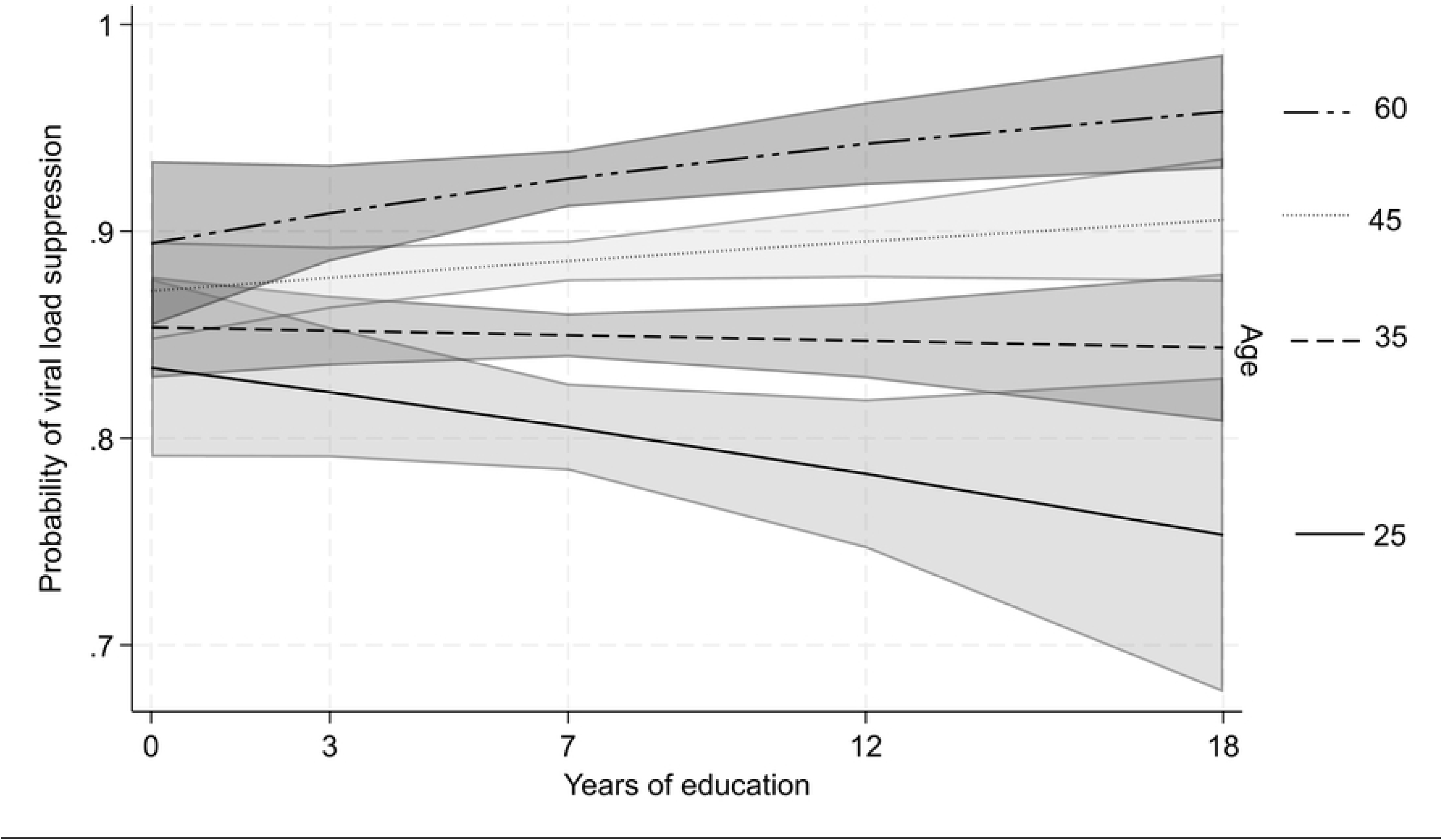
Probability of viral load suppression for each year of individual education among people living with HIV who initiated antiretroviral therapy with 95% confidence intervals, for ages 25, 35, 45, and 60 (from Table 4, Model 3).

## Discussion

Education is often cited as a mechanism to improve health and longevity in sub-Saharan Africa. However, there is limited and inconsistent evidence about whether formal education supports individuals’ capacity to manage their HIV treatment in the region. To fill this gap, I evaluated the role of education in facilitating HIV treatment adherence, operationalized as viral load suppression (or ‘viral load suppression’). I found no population-level associations between formal education and viral load suppression, and this was true for both men and women and for all age groups. A cursory takeaway from my findings might be that education is unimportant or at least not very important for HIV treatment adherence. However, I propose that this evidence offers deeper theoretical insights about heterogenous education-health relationships globally, and highlights key questions for future research about *why* formal education may not predict this type of chronic care management in the context of southern and eastern Africa.

Overall, these findings support my alternative supposition that specific contextual realities in the region may attenuate the pathways from education to effective treatment adherence. Throughout sub-Saharan Africa, there is concern among scholars, policy makers, and family members that, while more children have been attending school as a result of ‘Education for All’ policies, schooling is not leading to increased learning or employment (53). Education and health scholars have in turn hypothesized that these low returns from schooling may explain other inconsistent and paradoxical associations between education and health in the region (18). Here, I discuss four contextual factors that may explain the statistical independence between education and viral load suppression.

First, more education may not support PLWH with accessing greater material resources to overcome logistical barriers to HIV care and treatment. Consistent formal employment in the region is limited even for those with relatively greater schooling. Moreover, unlike contexts such as the United States, employer-sponsored private insurance is uncommon in most areas of sub-Saharan Africa and does not necessarily predict access to HIV services. Because HIV treatment is now free and available at most local clinics, the material resources required to access treatment on a routine basis are less burdensome compared to HIV treatment in the past and other areas of the world. In further support of these ideas, my unadjusted and adjusted analyses demonstrate that neither wealth nor employment were associated with viral load suppression. Given the dramatic reductions in HIV treatment provided by the U.S. government via the PEPFAR program in 2025—leading to reduced HIV treatment access—it will be important to reassess these associations over time.

Second, the lack of an association between education and viral load suppression may be explained by pervasive learning gaps in the region. Indeed, only 20-55% of school-aged children in these countries have obtained basic reading and math skills after completing 4^th^ or 5^th^ grade (33). Nevertheless, entering higher levels of secondary schooling (i.e., A levels) or tertiary education requires a clear demonstration of reading, writing, and mathematical proficiency. As such, I would have expected at least some additional benefit for treatment adherence when comparing those who continued their education beyond primary school with those who did not. Yet, even in my sensitivity analyses testing viral load suppression associations by education level (rather than years) there were no significant relationships observed (see Appendix 4).

Because literacy and numeracy measures are not available in the PHIA data, I cannot directly test the mediating role of such skills in supporting HIV treatment adherence. Nevertheless, my findings suggest several possibilities regarding literacy and numeracy skills. On the one hand, it is possible that such skills are simply not necessary for achieving viral load suppression in this context. On the other hand, such skills may indeed be critical for HIV care, but literacy and numeracy could be acquired through non-formal and informal education, as demonstrated in other contexts (54). A final possibility is that other, contextual factors may supersede the importance of these skills, attenuating any education-viral load suppression relationships. For example, if weather and road conditions prohibit clinic access—a frequent occurrence in rural areas (55)—even the most skilled and highly resourced individuals will find it difficult to refill their HIV treatment prescriptions.

Third, while self-efficacy, motivation, and risk-assessment are important for HIV treatment adherence, such factors can be obtained outside of formal schooling. Over the past decade, many myths and concerns about ARVs have been dispelled and many people know someone who has been helped by HIV treatment (22, 56). Many in the region are aware of the life-saving value of HIV treatment and the importance of treatment adherence (57). Thus, in the current era of Universal Treatment, education may be less important for facilitating individual’s knowledge about the importance of ART adherence for long-term health.

Fourth, other social forces—beyond education—may powerfully influence HIV treatment adherence, which could attenuate or complicate the relationship between education and viral load suppression in the region. It is difficult to overstate the positive impact of more widely available and tolerable HIV treatment regimens in the region. Literature from other settings demonstrates equitably designed and distributed health care systems can mitigate socio-economic disparities (58). At the time of this study, the more equitable structure of the HIV care system in these countries—namely free treatment and simpler regimens in urban and rural areas alike—may largely have attenuated many of the barriers that formal schooling would otherwise have helped individuals to overcome. Nevertheless, notable barriers to HIV care services and the complex demands of daily HIV treatment adherence persisted even after universal treatment expansions (59, 60). In the sample for my study, 14% of the population who had started ART remained virally unsuppressed—ample space for disparities to emerge. Indeed, my study alongside myriad others (13, 14), demonstrates clear disparities in viral load suppression by age and gender. The fact that education did not play a role in viral load suppression even within different age or cohort and gender groups suggests that either: a) other social forces beyond education-related material and cognitive skills—like gender norms and cohort or life stage/age differences—may simply be better predictors of viral load suppression disparities in the region; and/or b) other social forces (i.e., countervailing mechanisms (61))—like stigma or completing life priorities— may attenuate or complicate the importance of education-related skills and resources for viral load suppression in the region. Future research is needed to understand *why* there are no viral load suppression disparities by education when there are clearly other viral load suppression disparities in the region. In sum, my findings suggest that the causal relationship between education and health outcomes may be less fundamental and more contextually variable between lower and higher resource settings, deserving of additional theoretical and empirical examination globally.

The results I provide have important implications for policy. First, this study provides clarity about the relationship between formal education and HIV treatment adherence in southern and eastern Africa amidst prior inconsistent evidence (3, 62). Education is a valuable resource for many facets of life, and my findings should not be read as undermining the potential importance of education for health or society more broadly. Rather, this study contributes to the debate among international policy makers about the relevance, limits, and potential of formal schooling to improve chronic health outcomes in adulthood in the region (63). Second, a deeper understanding about the countervailing processes that may have attenuated the relationship between education and viral load suppression could inform future equity-focused chronic care interventions in the region. For example, cash transfer or women’s economic empowerment interventions are popular in the region for addressing a variety of social issues. Such investments may, in and of themselves, provide limited returns for addressing chronic HIV care outcomes (64, 65). Whereas innovative interventions may be needed that can specifically address why certain men and women, regardless of their education or wealth, struggle to adhere to their HIV medication.

### Strengths and limitations

The PHIA is the first study to consistently collect viral load and education measures at the population level across multiple countries in sub-Saharan Africa. Historically, viral load measures have been obtained through clinical data, which rarely includes education measures and does not include the population who are disengaged from HIV care. Using this novel data, I am able to examine education-viral load suppression associations in a consistent manner across countries. Further, in contrast to many studies that categorize education by level of completion, I measure educational continuously allowing for a me to examine the association within primary and secondary schooling, where the majority of the variance occurs in these contexts.

These findings should also be considered in light of several limitations. First, my use of a cross-sectional viral load measures fails to capture perfect adherence on a day-to-day basis—it is possible to have good, but imperfect adherence, and to still demonstrate viral load suppression on a blood test. Additionally, I am unable to measure sustained viral load suppression over time, limiting my ability to speak to the influence of education on successful, on-going maintenance in HIV care. Nevertheless, viral load suppression remains a critical indicator for individual and population health, and provides a clear marker of effective, if imperfect, treatment adherence at the population level. As my measure of education is based on self-report, rather than school records, responses may be influenced by recall or social desirability biases. Finally, it is possible that these findings are downwardly biased due to other unobserved factors or selection issues— for example, educational differences in HIV infection rates, entry into HIV care, or HIV-related mortality prior to the survey. Though, in separate analyses with this same population, I did not identify any relationship between educational attainment and HIV diagnosis or treatment initiation, which suggests that this form of selection bias will be less relevant for the present study (57).

## Conclusion

This study addresses important theoretical questions about education-health linkages globally and offers practical insights for chronic care interventions in sub-Saharan Africa. That education was not a predictor for viral load suppression provides a different perspective about the widely held belief that formal education facilitates health in the region. HIV care availability, the employment context, and low quality of education in the region may partially, but not fully, explain why education was not associated with HIV treatment adherence across these countries. More research is needed to understand the complex contextual factors and countervailing processes that may explain these surprising population-level findings.

## Data Availability

Data and code availability: All data is publicly available upon request from the PHIA study at https://phia-data.icap.columbia.edu/. All statistical code is available upon request from the author.

## Acknowledgements

I am grateful for the mentorship and feedback provided by Sara Yeatman, Patrick Krueger, Stephanie Robert, and Eric Grodsky as I developed this work. I want to thank all the respondents who shared their information with Population HIV Impact Assessment (PHIA) study, and the PHIA study teams for all their efforts to collect and share this data.

## Supporting Information

S1 Table. Sensitivity Analysis: Associations between education *levels* and viral load suppression (viral load suppression) among people living with HIV who initiated antiretroviral therapy, adjusted odds ratios (AOR) with 95% confidence intervals (CI)

S2 Table. Sensitivity Analysis: Multivariable associations between years of education and viral load suppression (viral load suppression) among people living with HIV who initiated antiretroviral therapy, adjusted odds ratios (AOR) with 95% confidence intervals (CI), by country

S3 Table. Sensitivity Analysis: Multivariable associations with viral load suppression with gender moderation, model 2, adjusted odds ratios (AOR) with 95% confidence intervals (CI), by country

S4 Table. Sensitivity Analysis: Multivariable associations with viral load suppression with age moderation, model 2, adjusted odds ratios (AOR) with 95% confidence intervals (CI), by country

## References

1. Jukes M, Simmons S, Bundy D. Education and vulnerability: the role of schools in protecting young women and girls from HIV in southern Africa. Aids. 2008;22:S41–S56.

2. Leon J, Baker DP, Salinas D, Henck A. Is education a risk factor or social vaccine against HIV/AIDS in sub-Saharan Africa? The effect of schooling across public health periods. Journal of Population Research. 2017;34(4):347–72.

3. Heestermans T, Browne JL, Aitken SC, Vervoort SC, Klipstein-Grobusch K. Determinants of adherence to antiretroviral therapy among HIV-positive adults in sub-Saharan Africa: a systematic review. BMJ global health. 2016;1(4):e000125.

4. Green D, Tordoff DM, Kharono B, Akullian A, Bershteyn A, Morrison M, et al. Evidence of sociodemographic heterogeneity across the HIV treatment cascade and progress towards 90-90-90 in sub-Saharan Africa–a systematic review and meta-analysis. Journal of the International AIDS Society. 2020;23(3):e25470.

5. Eisinger RW, Dieffenbach CW, Fauci AS. HIV viral load and transmissibility of HIV infection: undetectable equals untransmittable. Jama. 2019;321(5):451–2.

6. Nash D, Yotebieng M, Sohn AH. Treating all people living with HIV in sub-Saharan Africa: a new era calling for new approaches. Journal of Virus Eradication. 2018;4(Suppl 2):1.

7. Makofane K, van der Elst EM, Walimbwa J, Nemande S, Baral SD. From general to specific: moving past the general population in the HIV response across sub-Saharan Africa. Journal of the International AIDS Society. 2020;23:e25605.

8. Green D, Tordoff DM, Kharono B, Akullian A, Bershteyn A, Morrison M, et al. Evidence of sociodemographic heterogeneity across the HIV treatment cascade and progress towards 90-90-90 in sub-Saharan Africa - a systematic review and meta-analysis. J Int AIDS Soc. 2020;23(3):e25470.

9. Rabkin M, El-Sadr WM. Why reinvent the wheel? Leveraging the lessons of HIV scale-up to confront non-communicable diseases. Global public health. 2011;6(3):247–56.

10. Vorkoper S, Kupfer LE, Anand N, Patel P, Beecroft B, Tierney WM, et al. Building on the HIV chronic care platform to address noncommunicable diseases in sub-Saharan Africa: a research agenda. AIDS (London, England). 2018;32(Suppl 1):S107.

11. Beaglehole R, Epping-Jordan J, Patel V, Chopra M, Ebrahim S, Kidd M, et al. Improving the prevention and management of chronic disease in low-income and middle-income countries: a priority for primary health care. The Lancet. 2008;372(9642):940-9.

12. Muhoza DN. The heterogeneous effects of socioeconomic and cultural factors on fertility preferences: evidence from Rwanda and Kenya. Journal of Population Research. 2019;36(4):347–63.

13. Dovel K, Yeatman S, Watkins S, Poulin M. Men’s heightened risk of AIDS-related death: the legacy of gendered HIV testing and treatment strategies. AIDS (London, England). 2015;29(10):1123.

14. Inzaule SC, Kroeze S, Kityo CM, Siwale M, Akanmu S, Wellington M, et al. Long-term HIV treatment outcomes and associated factors in sub-Saharan Africa: Multicountry longitudinal cohort analysis. Aids. 2022;36(10):1437–47.

15. Lopus S, Frye M. Visualizing Africa’s Educational Gender Gap. Socius. 2018;4:2378023118795956.

16. LeVine RA, LeVine S, Schnell-Anzola B, Rowe ML, Dexter E. Literacy and mothering: How women’s schooling changes the lives of the world’s children: Oxford University Press; 2011.

17. Psaki SR, Chuang EK, Melnikas AJ, Wilson DB, Mensch BS. Causal effects of education on sexual and reproductive health in low and middle-income countries: A systematic review and meta-analysis. SSM-population health. 2019;8:100386.

18. Mensch BS, Chuang EK, Melnikas AJ, Psaki SR. Evidence for causal links between education and maternal and child health: systematic review. Tropical Medicine & International Health. 2019;24(5):504–22.

19. Bintabara D, Mwampagatwa I. Socioeconomic inequalities in maternal healthcare utilization: An analysis of the interaction between wealth status and education, a population-based surveys in Tanzania. PLOS Global Public Health. 2023;3(6):e0002006.

20. Gabrielli G, Paterno A, Salvini S, Corazziari I. Demographic trends in less and least developed countries: Convergence or divergence? Journal of Population Research. 2021;38(3):221–58.

21. Emily B. Zimmerman SHW, Amber Haley. Understanding the Relationship Between Education and Health: A Review of the Evidence and an Examination of Community Perspective.

22. Chamberlin S, Mphande M, Phiri K, Kalande P, Dovel K. How HIV Clients Find Their Way Back to the ART Clinic: A Qualitative Study of Disengagement and Re-engagement with HIV Care in Malawi. AIDS and Behavior. 2021:1–12.

23. Ware NC, Idoko J, Kaaya S, Biraro IA, Wyatt MA, Agbaji O, et al. Explaining adherence success in sub-Saharan Africa: an ethnographic study. PLoS Med. 2009;6(1):e1000011.

24. Rutherford ME, Mulholland K, Hill PC. How access to health care relates to under-five mortality in sub-Saharan Africa: systematic review. Tropical medicine & international health. 2010;15(5):508–19.

25. Lipkus IM, Peters E. Understanding the role of numeracy in health: Proposed theoretical framework and practical insights. Health Education & Behavior. 2009;36(6):1065–81.

26. Waldrop-Valverde D, Osborn CY, Rodriguez A, Rothman RL, Kumar M, Jones DL. Numeracy skills explain racial differences in HIV medication management. AIDS and Behavior. 2010;14(4):799–806.

27. Ashaba S, Baguma C, Tushemereirwe P, Nansera D, Maling S, Zanon BC, et al. Correlates of HIV treatment adherence self-efficacy among adolescents and young adults living with HIV in southwestern Uganda. PLOS Global Public Health. 2024;4(9):e0003600.

28. Rothman RL, Montori VM, Cherrington A, Pignone MP. Perspective: the role of numeracy in health care. Journal of health communication. 2008;13(6):583–95.

29. Izudi J, Cattamanchi A, Castelnuovo B, King R. Barriers and facilitators to viral load suppression among people living with HIV following intensive adherence counseling in Kampala, Uganda: A qualitative study. Social Science & Medicine. 2024;343:116595.

30. Pinheiro C, de-Carvalho-Leite J, Drachler M, Silveira V. Factors associated with adherence to antiretroviral therapy in HIV/AIDS patients: a cross-sectional study in Southern Brazil. Brazilian Journal of Medical and Biological Research. 2002;35(10):1173–81.

31. Mirowsky J, Ross CE. Education, learned effectiveness and health. London Review of Education. 2005;3(3):205–20.

32. McCullough EB. Labor productivity and employment gaps in Sub-Saharan Africa: The World Bank; 2015.

33. Declaration I, editor Education 2030: Towards inclusive and equitable quality education and lifelong learning for all. World Education Forum; 2015.

34. Goldman DP, Lakdawalla DN. A theory of health disparities and medical technology. Contributions in Economic Analysis & Policy. 2005;4(1).

35. Wei F, Mullooly JP, Goodman M, McCarty MC, Hanson AM, Crane B, et al. Identification and characteristics of vaccine refusers. BMC pediatrics. 2009;9:1–9.

36. Reich JA. “We are fierce, independent thinkers and intelligent”: Social capital and stigma management among mothers who refuse vaccines. Social Science & Medicine. 2020;257:112015.

37. Lutfey K, Freese J. Toward some fundamentals of fundamental causality: Socioeconomic status and health in the routine clinic visit for diabetes. American Journal of Sociology. 2005;110(5):1326–72.

38. UNESCO G. EFA Global Monitoring Report 2015: Education For All 2000-2015: Achievements and Challenges. Paris: UNESCO; 2015.

39. Desmond C, Watt K, Naicker S, Behrman J, Richter L. Girls’ schooling is important but insufficient to promote equality for boys and girls in childhood and across the life course. Development Policy Review. 2024;42(1):e12738.

40. Yeatman S, Chamberlin S, Dovel K. Women’s (health) work: A population-based, cross-sectional study of gender differences in time spent seeking health care in Malawi. PloS one. 2018;13(12):e0209586.

41. Heise L, Greene ME, Opper N, Stavropoulou M, Harper C, Nascimento M, et al. Gender inequality and restrictive gender norms: framing the challenges to health. The Lancet. 2019.

42. Maeri I, El Ayadi A, Getahun M, Charlebois E, Akatukwasa C, Tumwebaze D, et al. “How can I tell?” Consequences of HIV status disclosure among couples in eastern African communities in the context of an ongoing HIV “test-and-treat” trial. AIDS care. 2016;28(sup3):59–66.

43. Bohle LF, Dilger H, Groß U. HIV-serostatus disclosure in the context of free antiretroviral therapy and socio-economic dependency: experiences among women living with HIV in Tanzania. African Journal of AIDS Research. 2014;13(3):215–27.

44. Pritchett L. The rebirth of education: Schooling ain’t learning: CGD Books; 2013.

45. Frye M, Lopus S. From Privilege to Prevalence: Contextual Effects of Women’s Schooling on African Marital Timing. Demography. 2018;55(6):2371–94.

46. Assessments P-bHI. PHIA Data Use Manual. 2019.

47. Beretta L, Santaniello A. Nearest neighbor imputation algorithms: a critical evaluation. BMC medical informatics and decision making. 2016;16(3):197–208.

48. McMahon JH, Elliott JH, Bertagnolio S, Kubiak R, Jordan MR. Viral suppression after 12 months of antiretroviral therapy in low-and middle-income countries: a systematic review. Bulletin of the World Health Organization. 2013;91:377–85.

49. Labhardt ND, Bader J, Lejone TI, Ringera I, Hobbins MA, Fritz C, et al. Should viral load thresholds be lowered?: revisiting the WHO definition for virologic failure in patients on antiretroviral therapy in resource-limited settings. Medicine. 2016;95(28).

50. ICF. Demographic and Health Surveys Standard Recode Manual for DHS7. Rockville, Maryland, U.S.A.:; 2018.

51. Walsemann KM, Gee GC, Ro A. Educational attainment in the context of social inequality: new directions for research on education and health. American Behavioral Scientist. 2013;57(8):1082–104.

52. StataCorp. Stata Statistical Software: Release 18. College Station, TX: StataCorp LLC; 2023.

53. Kaler SWaA. Pivoting to Learning: A Puzzle with Many Pieces. 2016. Contract No.: RISE-WP-16/006.

54. Banerjee AV, Bhattacharjee S, Chattopadhyay R, Alejandro JG. The Untapped Math Skills of Working Children in India: Evidence, Possible Explanations, and Implications. August; 2017.

55. Merten S, Kenter E, McKenzie O, Musheke M, Ntalasha H, Martin-Hilber A. Patient-reported barriers and drivers of adherence to antiretrovirals in sub-Saharan Africa: a meta-ethnography. Tropical Medicine & International Health. 2010;15:16–33.

56. Ross J, Ingabire C, Umwiza F, Gasana J, Munyaneza A, Murenzi G, et al. How early is too early? Challenges in ART initiation and engaging in HIV care under Treat All in Rwanda—A qualitative study. PloS one. 2021;16(5):e0251645.

57. Chamberlin SA. Education and Chronic Care Management: The Case of HIV Care and Treatment in Southern and Eastern Africa [Ph.D.]. United States -- Colorado: University of Colorado at Denver; 2022.

58. Chang VW, Lauderdale DS. Fundamental cause theory, technological innovation, and health disparities: the case of cholesterol in the era of statins. Journal of health and social behavior. 2009;50(3):245–60.

59. Ware NC, Wyatt MA, Geng EH, Kaaya SF, Agbaji OO, Muyindike WR, et al. Toward an understanding of disengagement from HIV treatment and care in sub-Saharan Africa: a qualitative study. PLoS medicine. 2013;10(1):e1001369.

60. Comfort AB, Asiimwe S, Amaniyre G, Orrell C, Moody J, Musinguzi N, et al. Social networks and HIV treatment adherence among people with HIV initiating treatment in rural Uganda and peri-urban South Africa. SSM Popul Health. 2024;25:101593.

61. Freese J, Lutfey K. Fundamental Causality: Challenges of an Animating Concept for Medical Sociology. Handbook of the Sociology of Health, Illness, and Healing. Handbooks of Sociology and Social Research 2011. p. 67–81.

62. Chipanta D, Amo-Agyei S, Giovenco D, Estill J, Keiser O. Socioeconomic inequalities in the 90– 90–90 target, among people living with HIV in 12 sub-Saharan African countries—Implications for achieving the 95–95–95 target—Analysis of population-based surveys. EClinicalMedicine. 2022;53.

63. Chudgar A, Kim Y, Morley A, Sakamoto J. Association between completing secondary education and adulthood outcomes in Kenya, Nigeria, Tanzania and Uganda. International Journal of Educational Development. 2019;68:35–44.

64. Yotebieng M, Thirumurthy H, Moracco KE, Edmonds A, Tabala M, Kawende B, et al. Conditional Cash Transfers to Increase Retention in PMTCT Care, Antiretroviral Adherence, and Postpartum Virological Suppression: A Randomized Controlled Trial. JAIDS Journal of Acquired Immune Deficiency Syndromes. 2016;72:S124–S9.

65. Ayer A, Mukondwa RW, Aviles Guaman C, Takarinda K, West N, Makoni T, et al. Poverty and protection: the relationship between multidimensional poverty, social protections interventions, and HIV viral load. AIDS. 9900:10.1097/QAD.0000000000004276.

